# A Self-Administered Digital Test Battery for Early Detection of Cognitive Impairment in Alzheimer’s Disease

**DOI:** 10.1101/2025.07.16.25331196

**Authors:** Sophia Rutt, Marleen Taute, Paulina Tegethoff, Robert Perneczky, Carolin Kurz, Boris-Stephan Rauchmann

## Abstract

**Introduction:** Early detection of cognitive decline is crucial for timely intervention in Alzheimer’s disease (AD). Traditional paper-pencil assessments often face limitations in accessibility, lengthy administration, and interrater reliability, restricting their broader application across diverse populations. Digital tools may overcome these restrictions by offering user-friendly, non-verbal, and scalable assessments. This study investigated the diagnostic accuracy of BraincheX, a newly developed digital test battery designed with a focus on accessibility and usability, by comparing its subtest performance to equivalents of the Consortium to Establish a Registry for AD (CERAD)-Plus battery.

**Methods:** Fifty-six participants with early AD and healthy controls were recruited from the LMU hospital memory clinic. Diagnostic groups were defined by amyloid positivity and clinical expert consensus. Participants completed both the BraincheX digital battery and the CERAD-Plus assessment. A total BraincheX score was calculated by summing standardized subtest scores. Data were analyzed using receiver operating curve (ROC) analysis and the Youden Index to determine diagnostic accuracy. Correlations between BraincheX and CERAD-Plus subtests were examined, and group differences were analyzed via ANOVA.

**Results:** ROC analysis revealed strong diagnostic accuracy for the BraincheX total score in differentiating between early AD and controls (AUC = .86; optimal cut-off = -1.51), with a sensitivity of 89.7% and a specificity of 32%. BraincheX total scores correlated strongly with CERAD total scores (r = .73, p < .001). BraincheX demonstrated high internal consistency (Cronbach’s α = .84). ANOVA confirmed significant group differences across all BraincheX subtests (p < .01). Additionally, individual BraincheX subtests showed strong correlations with their CERAD-Plus equivalents, such as the Clock Drawing Test (r = .73) and TMT-B (r = .48).

**Discussion:** These findings support BraincheX as a promising, accessible digital screening tool for cognitive decline, offering advantages such as automated scoring, immediate feedback, and reduced examiner bias. Its design promotes greater equity and inclusivity in cognitive assessment. Future research is underway to validate BraincheX in larger and more diverse populations.

## 1 Introduction

Early assessment and diagnosis of cognitive decline are essential for the effective clinical management of Alzheimer’s disease (AD), as neuropathological changes often precede symptoms by years [18]. Early diagnosis empowers individuals to actively manage their health, make informed decisions, and engage in personalized preventive strategies, aiming to slow disease progression. It also opens opportunities to maintain autonomy, build resilience, and preserve quality of life for as long as possible [9].

Standardized cognitive assessments, such as the Consortium to Establish a Registry for Alzheimer’s Disease (CERAD)-Plus test battery, the Montreal Cognitive Assessment (MoCA), and the Mini-Mental-State Examination (MMSE) were developed for rater-administered cognitive screenings, limiting their more widespread use. Such limitations include the absence of remote administration capabilities, restricted potential for longitudinal assessment, lack of integration of multimodal data, and reliance on trained raters, which introduces potential for rater bias and variability in administration and scoring [14].

Most traditional paper-and-pencil cognitive instruments tend to be more sensitive to deficits in later disease stages and are typically administered in specialized clinical settings, which can limit their utility for early detection and broad accessibility. More recent tools like the MoCA offer improved sensitivity for earlier stages but still rely on trained personnel and structured environments [14]. In Germany as in many other industry nations, waiting times for AD diagnostic assessments are long, with an estimated delay of 50 months for diagnostic evaluation and determination of eligibility for disease-modifying treatments from 2024 to 2043 [22]. Barriers to traditional dementia diagnostics include limited access to diagnostic services, a lack of awareness regarding non-memory cognitive impairments, and the complexity of the clinical diagnostic process [10] [14]. With the introduction of disease-modifying treatments for early-stage AD, the need for faster and more efficient clinical decision-making is becoming increasingly urgent to avoid delays in diagnosis and prevent unnecessary disease progression [22].

Digital cognitive test batteries offer a promising alternative to paper-and-pencil methods by enabling precise measurement of response times, standardized stimulus presentation, and automated comparison to individual baselines and normative data [3] [10] [14]. They are more efficient and cost-effective, often self-administered, and require minimal rater training, thereby reducing examiner bias through standardized procedures [14]. In addition, digital tools capture nuanced behavioral data, such as spatial planning strategies, and provide immediate, analyzable feedback for clinicians and participants [3] [6] [14].

In this study we assessed the performance in differentiating early AD individuals from healthy controls of BraincheX (Medotrax GmbH, Munich, Germany), a self-administered novel digital test battery designed to evaluate cognitive function across multiple domains, including cognitive flexibility, processing speed, visual and spatial memory, abstraction, and planning skills. BraincheX offers a non-verbal format that minimizes the impact of language barriers and integrates digital variants of traditional tests alongside a newly developed visual memory test. Through standardized administration and automated scoring, it reduces examiner bias and captures data relevant to everyday functioning.

## 2 Methods

### 2.1. Participants

Participants were recruited consecutively from the memory clinic at the Department of Psychiatry and Psychotherapy, LMU Hospital, Ludwig-Maximilians-University Munich, between October 2024 and February 2025. All individuals presented for a standard diagnostic work-up due to memory complaints or suspected cognitive impairment. A total of 56 participants meeting the inclusion criteria were enrolled and completed both the BraincheX and CERAD-Plus test batteries. Inclusion criteria encompass the following: (1) referral for cognitive evaluation, (2) provision of signed, written, and dated informed consent, (3) confirmed capacity to consent, (4) age of 50 years or older, and (5) expert diagnosis of early AD according to the German S3 Dementia Guideline (2023) — classified as mild cognitive impairment (MCI) or mild dementia due to AD — or as an age-matched cognitively healthy control. MCI was defined by objective cognitive deficits without significant impairment of daily functioning. Early AD diagnosis adhered to the IWG-2 criteria, requiring both clinical phenotype and biomarker evidence (amyloid positivity via cerebrospinal fluid [CSF] analysis and/or positron emission tomography [PET]). Exclusion criteria included (1) fewer than nine years of formal education, (2) diagnosis of any neurodegenerative disease other than AD, and (3) moderate to severe dementia stages. Diagnostic classification was based on clinical expert consensus incorporating biomarker profiles [18] when available (n=18). For the following analysis, participants were categorized into two groups: amyloid-positive early AD (n=25; mild AD dementia n=12; MCI-AD n=13) and amyloid-negative cognitively healthy controls (n=31). Participants were contacted via telephone and recruited through convenience sampling. Travel expenses were reimbursed. All participants were fully informed about the study procedures and provided written informed consent. The study was approved by the Ethics Committee of LMU Munich (project number 22-1117) and is registered at ClinicalTrials.gov (NCT06711952).

### 2.2 Evaluation of the digital neurocognitive Test Battery BraincheX

BraincheX is a mobile application for cognitive assessment, developed by Medotrax GmbH, Munich, Germany, offering digital variations of established paper-and-pencil tests alongside a newly developed visual memory task. It is designed for efficient, accessible cognitive screening, with an average completion time of 23.60 minutes depending on the participant’s cognitive state.

The BraincheX battery includes:

- A modified **Symbol Digit Test** (SZT), similar to the RBANS and WAIS-IV subtests, presenting one practice example followed by assignments across 9 symbols within a 60-second time limit. Performance is scored as the number of correct responses, with adapted normative references based on age groups [11] [26].
- A **Visual Memory Test (VMT)**, presenting 20 images for encoding and subsequently 35 images (mix of new and familiar), requiring recognition decisions. The task structure is unique, though comparable to aspects of the Wechsler Memory Scale (WMS-IV Visual Reproduction) and the Visual Paired Comparison (VPC) tasks [38]. Normative data were acquired within the current project, as no directly equivalent test existed previously. The structure of the VMT and additional memory recognition tasks mimic the word recognition paradigms used in the CERAD, incorporating principles of discriminability without the language dependence [5] [28] [34] [35] [38].
- A **Line Orientation Test** similar to the RBANS structure, where 10 pairs of lines must be matched for orientation within 15 seconds per item. Scores are compared against adjusted norms to account for the added time constraint [11] [26].
- A digital **Pathfinding Test** comparable with the Trail Making Test Part B (TMT-B), in which participants are required to alternately connect 13 numbers and letters in ascending order. Performance is measured by time to completion with a maximum allowed duration of 300 seconds. Trials exceeding this limit are automatically capped [1] [28] [37|.
- A digital version of the **Clock Drawing Test** rated by using the Shulman’s procedure, requiring participants to depict “10 past 11” on a pre-drawn circle following detailed standardized instructions [34].

As a reference standard, the CERAD-Plus battery was used, comprising tests for verbal fluency, naming, memory encoding and recall, recognition memory, visuoconstruction, and executive function (Table 1). Subtests include Animal Naming, Phonemic Fluency, Trail Making Test A and B, Word List Learning, Recall and Recognition, Figure Recall, Modified Boston Naming Test, and Figure Copy. CERAD-Plus requires approximately 30 to 40 minutes to administer and is widely applied in clinical and research contexts for detecting and tracking cognitive decline. [24] [31].

**Table 1.** Comparison CERAD-Plus vs. BraincheX.

| Feature | CERAD-Plus | BraincheX |
| --- | --- | --- |
|  | Paper-pencil | Fully digital |
| Domains assessed | Memory, language, visuoconstruction, executive functions | Memory (visual & verbal), processing speed, cognitive flexibility, abstraction |
| Test-taking Time | 30 to 40 minutes | 15 to 25 minutes |
| Administration | Examiner-administered | Self-administered with minimal supervision |
| Scoring | Manual scoring, prone to inter-rater variability | Immediate results calculation |
| Language dependence | Language-heavy | Language-independent (e.g., visual memory test) |
| Setting | Specialized memory clinics | Scalable and broader, decentralized deployment |

*Note.* This table compares the features of the CERAD-Plus and BraincheX test battery regarding Domains assessed, Test-taking time, Administration, Scoring, Language dependence, Ecological validity, Setting

**Table 2.** Descriptive Statistics of the Study Population (N = 56)

| Variable | Category | Frequency | Percentage (%) |
| --- | --- | --- | --- |
| Diagnosis |  |  |  |
| Early AD |  | 25 | 44.6 |
| Control |  | 31 | 55.4 |
| Age | Range (years) | 50–87 | - |
| Education |  |  |  |
| Primary |  | 11 | 19.6 |
| Secondary |  | 24 | 42.9 |
| Tertiary |  | 21 | 37.5 |
| Gender |  |  |  |
| Female |  | 37 | 66.1 |
| Male |  | 19 | 33.9 |
| Marital Status |  |  |  |
| Married |  | 31 | 55.4 |
| Divorced |  | 12 | 21.4 |
| Single |  | 5 | 8.9 |
| Widowed |  | 6 | 10.7 |
| Domestic Partnership |  | 2 | 3.6 |
| Occupational Status |  |  |  |
| Retired |  | 39 | 69.6 |
| Occupied (employee) |  | 8 | 14.3 |
| Unoccupied (home-makers) |  | 3 | 5.4 |
| Other |  | 3 | 5.4 |
| Disability pensioner |  | 3 | 5.4 |

*Note.* This table demonstrates the demographic characteristics of the study population. All *p*-values are based on two-tailed tests. No significant difference in age between the early AD and control group ( $p = .19$ ). No significant difference in education between the early AD and control group ( $p = .27$ ). No significant difference in sex between the early AD and control group ( $p = .39$ ). A significant difference was found in marital status between early AD and control group ( $p = .05$ ). A significant difference was found in occupation between early AD and control group ( $p = .02$ ).

### 2.1 Study Design

The aim of this prospective, non-interventional, cross-sectional pilot study was to compare the performance and usability of the BraincheX and CERAD-Plus test batteries. Participants meeting the inclusion criteria were recruited to complete the BraincheX and CERAD-Plus test battery. For comparison, CERAD-Plus paper-based assessments conducted in the same patients within an average interval of 4.9 months before the BraincheX testing served as the reference standard [6].

The BraincheX test battery was administered in a controlled setting, with a trained psychologist present to assist if necessary and to ensure standardization across participants. Participants completed the BraincheX test battery in a quiet environment. An Apple iPad (iOS 17.5.1) and Apple Pencil Pro were provided to ensure standardized input conditions. Participants were instructed to carefully read the on-screen instructions and could ask the examiner if questions arose. Sociodemographic information such as age, sex, years of education and occupation were obtained. Test results were automatically recorded and processed by the BraincheX application. Data from the CERAD-Plus test battery were obtained from clinical records or prior research participation. These assessments had been administered by trained professionals in clinical or research settings according to standard protocols.

#### 2.4.1 Statistical Analysis

Pseudonymized data from BraincheX and paper-based questionnaires were analyzed using IBM SPSS Statistics [16]. Due to missing data, two participants were excluded (final N = 54). Descriptive characteristics (education, age, marital status, diagnosis) were calculated for initial group comparisons between AD and controls. Following the principles of CERAD-Plus total score calculation, BraincheX also calculates a total score (BraincheX score). For CERAD-Plus, six core subtests (Animal Naming, Boston Naming, Word List Learning, Recall, Recognition, Figure Copy) were included, excluding domains primarily relevant for Parkinson’s disease differentiation (e.g., TMT-A/B, phonemic fluency, figure recall) [7] [21] [39]. The BraincheX score is calculated as the sum of Z-scores of all previously described subtests, with the TMT-B completion time and Clock Drawing Test score inverted prior to summation. Z-scores are computed based on control group normative data (N = 29), corrected for age, sex, and education, and summed across five tests (SZT, VMT, LOT, TMT-B, Clock Drawing Test) [34]. Internal consistency was evaluated using Cronbach’s α. Diagnostic accuracy was assessed via receiver operator (ROC) curve analyses for each subtest and the BraincheX score. Study specific cutoffs were determined using the Youden Index. Correlations between corresponding BraincheX and CERAD-Plus subscores were analyzed, and differences between diagnostic groups were tested with univariate ANOVA adjusted for age, sex and education and post-hoc comparisons.

## 3 Results

### 3.1 Demographic characteristics

The study cohort included 21.4% of participants diagnosed with AD dementia (n=12), 23.2% with MCI (n=13), and 55.3% (n=31) were cognitively healthy controls. This distribution reflects a balanced representation of individuals across the continuum of cognitive functioning, providing a suitable sample for evaluating the diagnostic accuracy of BraincheX compared to traditional assessments. The final sample consisted of 56 participants aged between 50 and 87 years. The majority of the cohort was female (66.1%) and most participants (69.6%) were retired, 14.3% were actively employed, and smaller proportions were homemakers (5.4%), disability pensioners (5.4%), or self-occupied/other (5.4%).

### 3.2. Performance of the Control Group on BraincheX

The average completion time in minutes for the BraincheX test battery was *M* = 23.60, *SD* = 11.50, *range* = 11 – 68 minutes. The normative data of BraincheX subtests based on the control group is shown in Table 1.

### 3.4. Group Comparisons

Group differences in BraincheX subtest performance were analyzed using univariate ANOVAs (Figures 2). Significant differences between diagnostic groups were observed across all subtests: Symbol Digit Test (SZT; *F*(1,54) = 17.60 *p* < .001), Visual Memory Test (VMT; *F*(1,52) = 16.70, *p* < .01), Line Orientation Test (LOT; *F* (1,54) = 11.42, *p* < .01.003), Trail Making Test B (TMT-B; *F* (1,54) = 30.80, *p* < .001), and Clock Drawing Test (*F* (1,54) = 14.19, *p* < .001). Education consistently showed a significant main effect across all subtests, whereas sex had no significant influence (*p* > .10). Age was only significantly associated with performance on the SZT (*p* < .05). Assumptions of homogeneity of variance were evaluated using Levene’s test. While this assumption was met for most subtests, violations were detected for the VMT and TMT-B (*p* < .05). Accordingly, Welch’s ANOVA was conducted for these subtests, which confirmed significant group effects for the LOT (*F* (1,54) = 12.61, *p* < .001, *η*^*2*^ = .19) and TMTB (*F* (1,52) = 33.94, *p* < .001, *η*^*2*^ = .40). These results are depicted in Figure 2 and 3.

**Figure 2.**
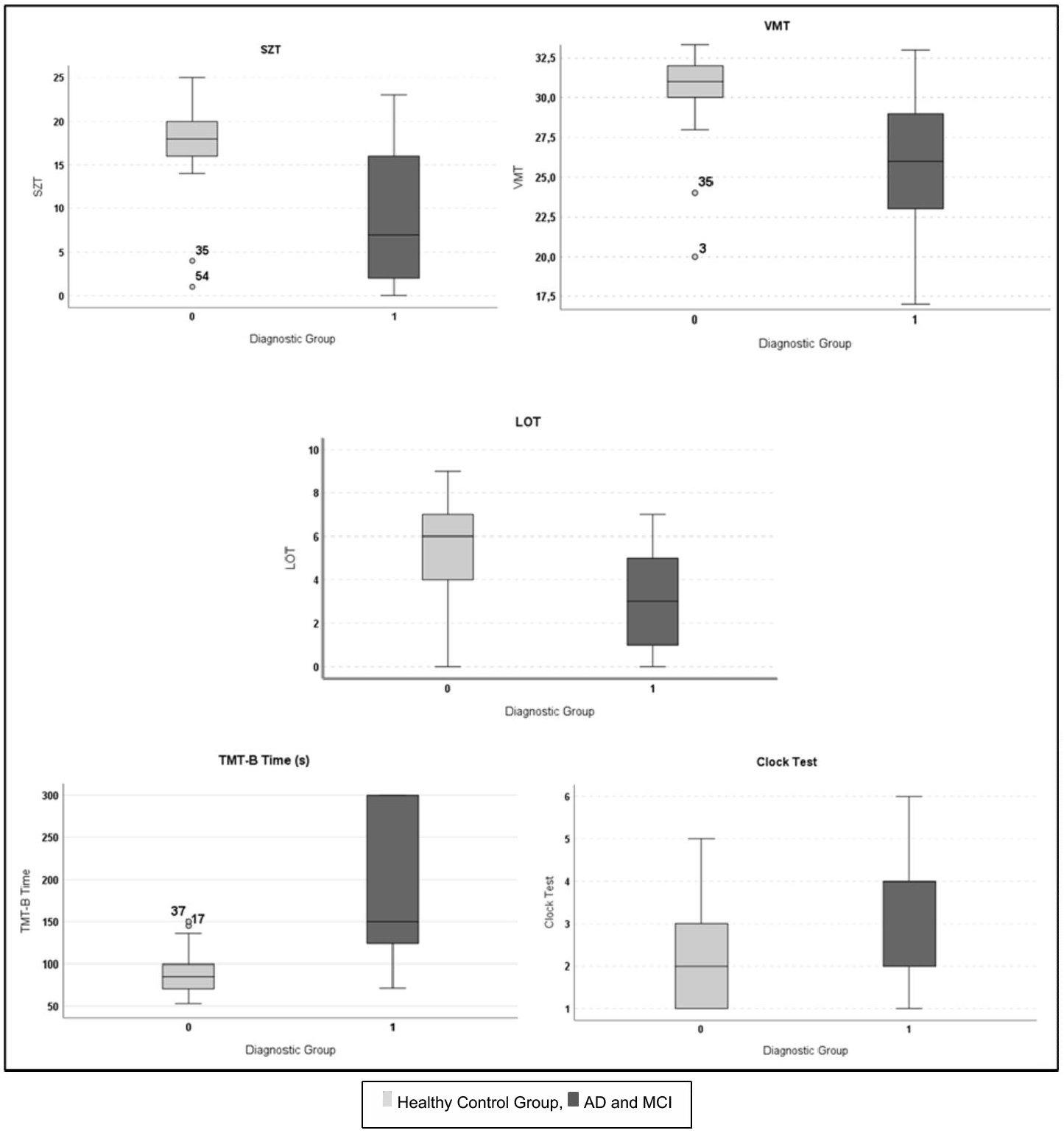
Comparison of Diagnostic Groups based on BraincheX Subtests. *Note*. Diagnostic group differences in BraincheX Subtests (SZT, VMT, LOT, Clock Drawing Test and TMT-B).

### 3.3. Psychometric Properties and Diagnostic Utility of BraincheX

To assess the internal consistency of BraincheX, Cronbach’s α was calculated, yielding a high reliability coefficient (*Cronbach’s α* = .84) [15]. Diagnostic accuracy was evaluated through receiver operating characteristic (ROC) curve analysis, resulting in an area under the curve (*AUC*) of .86, indicating strong discriminative power between early AD and healthy controls. The optimal cut-off score was determined using the Youden Index, identifying a BraincheX total score threshold of -1.51. This cut-off maximized sensitivity and specificity, achieving 89.7% sensitivity and 32% specificity (see Table 4 and Figure 4).

**Figure 4.**
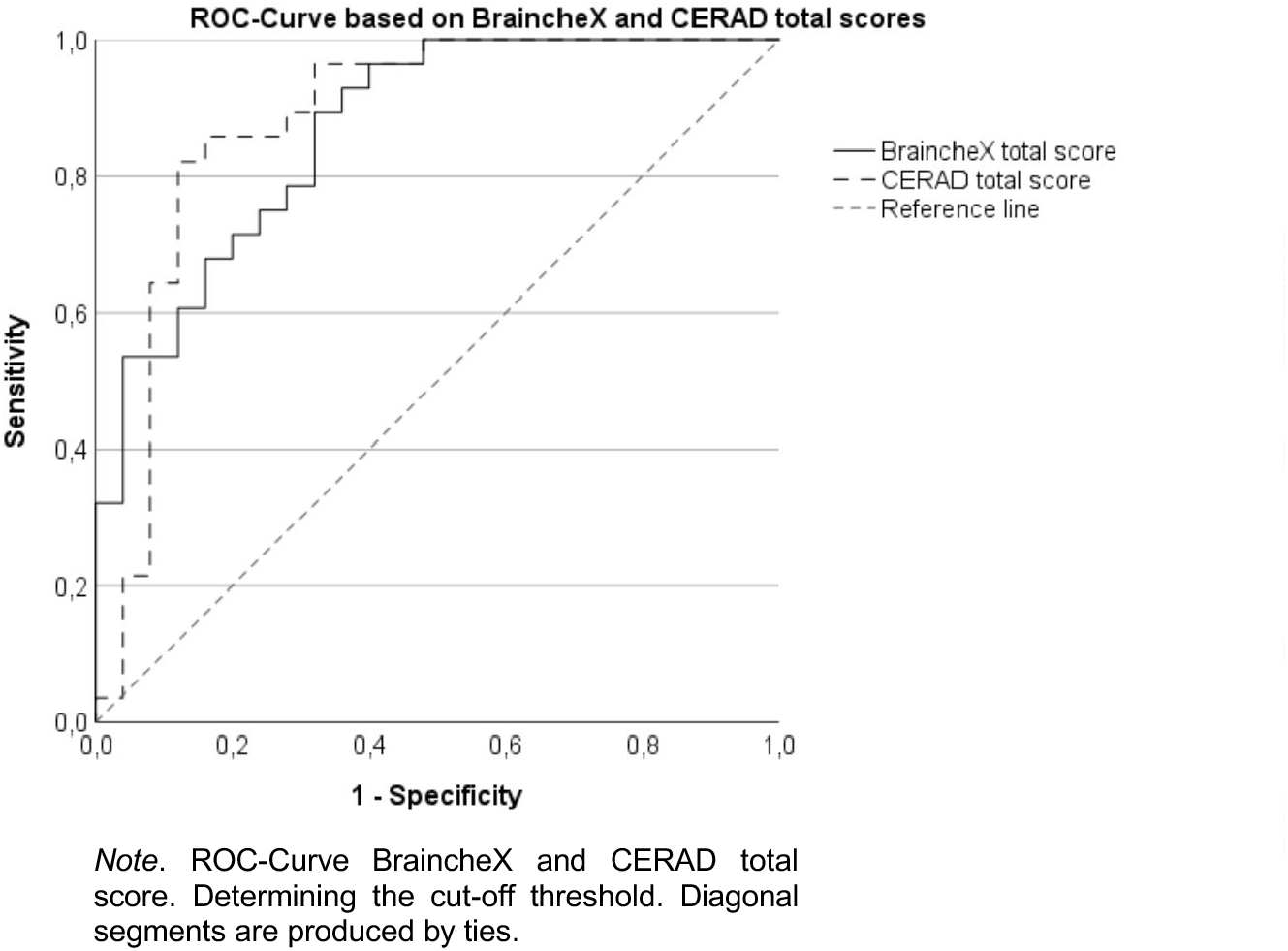
*ROC*-*Curve based on BraincheX and CERAD Total Score*.

### 3.5 BraincheX and CERAD-Plus statistical Comparisons

The convergent validity of the BraincheX total score with the CERAD Plus total score was examined using partial correlations. A strong positive correlation was found (*r* = .73, *p* < .001), suggesting that BraincheX measures cognitive domains closely related to those assessed by established paper-and-pencil batteries.

Associations between BraincheX and corresponding CERAD-Plus subtests were examined using partial correlation analyses, controlling for age, sex, and education. Significant positive correlations were observed between the BraincheX and CERAD Clock Drawing Tests (*r* = .47, *p* < .05), TMT-B times (*r* = .44, *p* < .05), and figure construction performance (*r* = .46, *p* < .05). Furthermore, the BraincheX Visual Memory Test (VMT) showed positive associations with several CERAD verbal memory subtests: Word List Recall (*r* = .36, *p* < .05), Animal Naming (*r* = .50, *p* < .01) and Boston Naming (*r* = .49, *p* < .01). These findings are visualized in table 3, illustrating the strength of associations between specific BraincheX and CERAD-Plus cognitive domains.

**Table 3.**
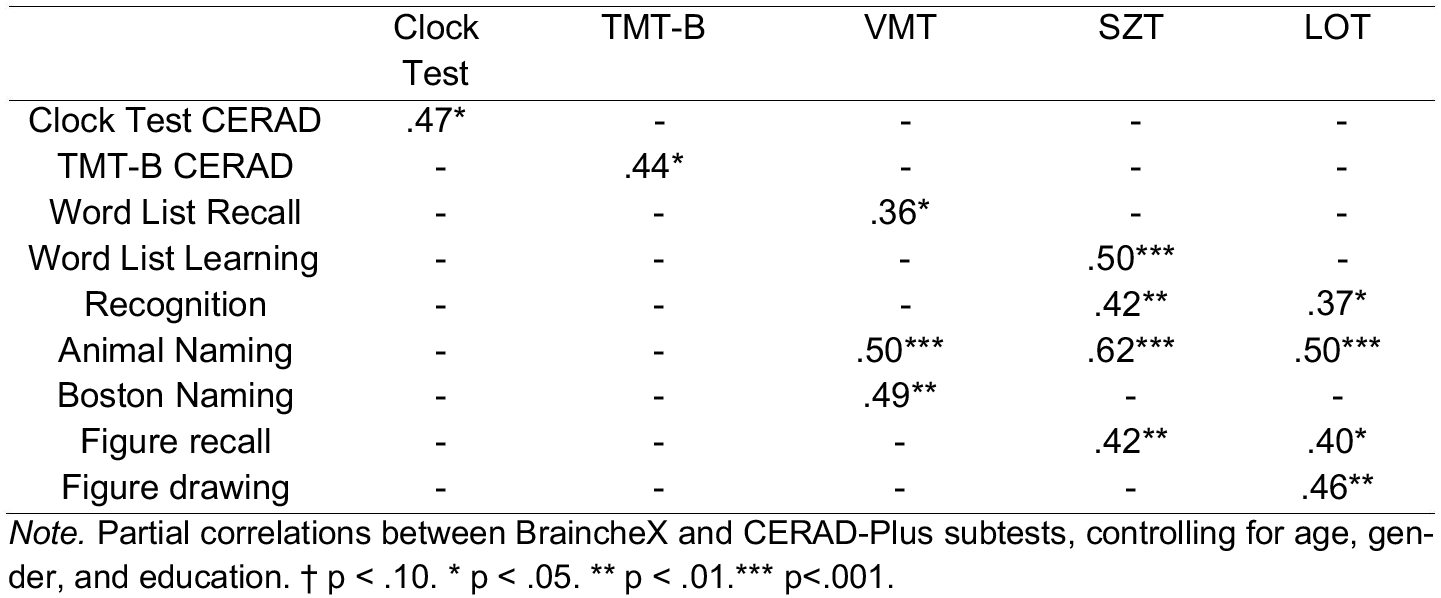
Partial Correlations controlled for age, sex and education.

|  | Clock<br>Test | TMT-B | VMT | SZT | LOT |
| --- | --- | --- | --- | --- | --- |
| Clock Test CERAD | .47* | - | - | - | - |
| TMT-B CERAD | - | .44* | - | - | - |
| Word List Recall | - | - | .36* | - | - |
| Word List Learning | - | - | - | .50*** | - |
| Recognition | - | - | - | .42** | .37* |
| Animal Naming | - | - | .50*** | .62*** | .50*** |
| Boston Naming | - | - | .49** | - | - |
| Figure recall | - | - | - | .42** | .40* |
| Figure drawing | - | - | - | - | .46** |

*Note.* Partial correlations between BraincheX and CERAD-Plus subtests, controlling for age, gender, and education. † $p < .10$ . \* $p < .05$ . \*\* $p < .01$ . \*\*\* $p < .001$ .

## 4 Discussion

Reliable and valid digital cognitive assessments are critical to enable early and accurate diagnosis of AD, especially as disease-modifying treatments emerge. In this pilot study, BraincheX demonstrated strong psychometric properties, including robust discriminative power between early AD and healthy controls (*AUC* = .86) with high internal consistency (*Cronbach’s α* = .84), and strong convergent validity with the CERAD-Plus total score (*r* = .73, *p* < .001). These results support BraincheX as a promising digital alternative to traditional assessments, offering benefits in efficiency and accessibility. BraincheX’s self-administration and automated scoring eliminate the need for an experienced examiner, reduce administrative time and enhance objectivity through automated scoring. This streamlines the clinical workflow and enhances scalability in research and real-world settings. The digital format reduces rater bias, provides immediate results, improving reliability of test results across experimenters and settings. These features position BraincheX as an efficient and scalable assessment toll in the early detection of AD.

The subtest correlation analysis supports the coverage if cognitive subdomains of BraincheX. Moderate to strong associations between executive tasks (e.g., TMT-B, Clock Test) and memory-related CERAD subtests (e.g., Word List Learning, Recognition) suggest overlapping constructs. Language-related subtests, such as Animal Naming and Boston Naming, were significantly associated with both memory tasks. Similarly, correlations with visuospatial subtests (e.g., Figure Recall, Figure Drawing) further demonstrate that BraincheX covers multiple cognitive subdomains relevant to early AD pathology. These findings underscore the battery’s construct validity and its ability to capture a multidimensional cognitive profile in a brief, digital format. Our findings align with previous research demonstrating that digital cognitive assessments can match conventional paper-based diagnostics in detecting MCI and dementia [8] [25]. In particular, BraincheX’s non-verbal and self-administered format addresses language and cultural barriers, supporting inclusivity across diverse populations. Furthermore, the digital provision facilitates access in underserved or remote areas, enabling earlier cognitive monitoring and intervention. BraincheX’s ability to deliver immediate feedback also positions it as a valuable tool for longitudinal cognitive tracking in clinical practice. Additionally, BraincheX may help to identify patients suitable for disease modifying treatments more efficiently. The expected implications of disease modifying treatments foresee, that there will be a significant change in need of diagnostic assessments [4]. BraincheX could contribute to streamlining both patient management and care pathways overall. As a result, modified treatment can become more accessible.

Despite these advantages, ethical concerns must be carefully managed, particularly regarding the automated delivery of potentially distressing results [30] [40]. Structured communication protocols and clinical follow-up by trained clinicians are essential to avoid anxiety and misinterpretation. Additionally, BraincheX’s integration into clinical workflows must be clearly defined, emphasizing its role as a tool for detecting signs of cognitive impairment—not as a standalone diagnostic instrument. Clinical diagnosis of AD should be based on a comprehensive assessment that includes BraincheX alongside other clinical evaluations, imaging, laboratory tests, and patient history.

Disparities in digital literacy remain a challenge [32] [27]. Some participants expressed hesitation or declined participation due to low confidence with digital tools, often linked to limited technological exposure or age-related decline [12] [32] [33]. Future research will systematically assess usability, digital affinity, and subjective workload to identify barriers and optimize user-centered design.

Several study specific limitations must be acknowledged. The relatively small sample size and the mean time interval of 4.91 months between CERAD-Plus and BraincheX assessments may have influenced the results. Nonetheless, previous research suggests that such intervals have limited impact on memory performance [20]. Future studies will aim for larger, more diverse cohorts and minimize the time gap between assessments to increase comparability. Moreover, validating BraincheX across a broader range of cognitive disorders—such as Parkinson’s disease, stroke, and depression—could substantially enhance its clinical applicability and utility.

## 5 Conclusion

This study demonstrates BraincheX as an efficient and accessible digital tool for detecting cognitive impairment, particularly in Alzheimer’s disease. Its high internal consistency and strong convergent validity with the CERAD-Plus total score support its role as a digital alternative to traditional assessments. Automated administration and scoring enhance clinical consistency and scalability, while the non-verbal, culturally neutral format increases accessibility.

## 6 Declarations

### 6.3 Ethics Approval

The study was approved by the Ethics Committee of Ludwig-Maximilians-University Munich (project number 22-1117) and conducted in accordance with the Declaration of Helsinki. Participants were recruited via telephone through convenience sampling and provided written informed consent prior to participation. Travel expenses were reimbursed. The study is registered at ClinicalTrials.gov (NCT06711952). Permission to reproduce material from other sources was not required, as only proprietary data were used.

### 6.4 Consent to Participate

All authors confirm that the study adhered to applicable ethical standards and legal requirements of the country in which the research was conducted. Written informed consent was obtained from all participants in accordance with the Declaration of Helsinki (1964).

### 6.5 Data Availability

The datasets generated and/or analyzed during the current study are available from the corresponding author on reasonable request.

## 6.6 Acknowledgments

We sincerely thank all study participants and their families for their valuable time, trust, and contribution to this research.

## 6.7 Authors Contributions

Sophia Rutt: Study conceptualization and Methodology, Data acquisition, Data analysis and interpretation, Original Draft writing and critical review.

Marleen Taute: Study conceptualization and Methodology, Data acquisition, Data analysis and interpretation, Review and Editing.

Paulina Tegethoff: Study conceptualization and Methodology, Data analysis and interpretation, Review and Editing.

Boris-Stephan Rauchmann: Study conceptualization and methodology, Data acquisition, Data analysis and interpretation, Original Draft writing and critical review, Approval of final version for Submission.

Carolin Kurz: Conceptualization and design of the study; development of methodology and digital adaptation procedures; primary data analysis and interpretation; major role in drafting, reviewing, and editing the manuscript; approval of the final version for submission.

Robert Perneczky: Study conceptualization and methodology, Data acquisition, Data analysis and interpretation, Original Draft writing and critical review, Approval of final version for Submission.

### 6.8 Open Access Statement

This article is distributed under the terms of the Creative Commons Attribution 4.0 International License (http://creativecommons.org/licenses/by/4.0/), which permits unrestricted use, distribution, and reproduction in any medium, provided the original author(s) and the source are credited.

## References

1. Ashendorf, L., Jefferson, A. L., O’Connor, M. K., Chaisson, C., Green, R. C., & Stern, R. A. (2008). Trail Making Test errors in normal aging, mild cognitive impairment, and dementia. Archives of clinical neuropsychology : the official journal of the National Academy of Neuropsychologists, 23(2), 129–137. 10.1016/j.acn.2007.11.005

2. Atkinson, R. C., & Shiffrin, R. M. (1971). The control of short-term memory. Scientific American, 225, 82–90.

3. Bauer, R. M., Iverson, G. L., Cernich, A. N., Binder, L. M., Ruff, R. M., & Naugle, R. I. (2012). Computerized Neuropsychological Assessment Devices: Joint Position Paper of the American Academy of Clinical Neuropsychology and the National Academy of Neuropsychology. The Clinical Neuropsychologist, 26(2), 177–196. 10.1080/13854046.2012.663001

4. Belder, C. R. S., Schott, J. M., & Fox, N. C. (2023). Preparing for disease-modifying therapies in Alzheimer’s disease. The Lancet. Neurology, 22(9), 782–783. 10.1016/S1474-4422(23)00274-0

5. Benton AL, Sivan A, Hamsher K, Varney N, Spreen O. Contributions to Neuropsychology Assessment: A Clinical Manual. 2. New York: Oxford University Press; 1994.

6. Berres M, Monsch AU, Bernasconi F, Thalmann B, Stähelin HB. Normal ranges of neuropsychological tests for the diagnosis of Alzheimer’s disease. Stud Health Technol Inform. 2000; 77:195-9. PMID: 11187541.

7. Chandler MJ, Lacritz LH, Hynan LS, Barnard HD, Allen G, Deschner M, Weiner MF, Cullum CM. A total score for the CERAD neuropsychological battery. Neurology. 2005 Jul 12;65 (1): 102–6.doi: 10.1212/01.wnl.0000167607.63000.38. PMID: 16009893.

8. Chan, J. Y. C., Yau, S. T. Y., Kwok, T. C. Y., & Tsoi, K. K. F. (2021). Diagnostic performance of digital cognitive tests for the identification of MCI and dementia: A systematic review. Ageing research reviews, 72, 101506. 10.1016/j.arr.2021.101506

9. Cummings, J., Apostolova, L., Rabinovici, G. D., Atri, A., Aisen, P., Greenberg, S., Hendrix, S., Selkoe, D., Weiner, M., Petersen, R. C., & Salloway, S. (2023). Lecanemab: Appropriate Use Recommendations. The journal of prevention of Alzheimer’s disease.., 10(3), 362–377. 10.14283/jpad.2023.30

10. Ding Z, Lee TL, Chan AS. Digital Cognitive Biomarker for Mild Cognitive Impairments and Dementia: A Systematic Review. J Clin Med. 2022 Jul 19;11(14):4191. doi: 10.3390/jcm11144191. PMID: 35887956; PMCID: PMC9320101.

11. Duff, K., Humphreys Clark, J. D., O’Bryant, S. E., Mold, J. W., Schiffer, R. B., & Sutker, P. B. (2008). Utility of the RBANS in detecting cognitive impairment associated with Alzheimer’s disease: sensitivity, specificity, and positive and negative predictive powers. Archives of clinical neuropsychology : the official journal of the National Academy of Neuropsychologists, 23(5), 603–612. 10.1016/j.acn.2008.06.004

12. Fischer, S., & Puschmann, C. (2021). Wie Deutschland über Algorithmen schreibt: Eine Analyse des Mediendiskurses über Algorithmen und Künstliche Intelligenz (2005–2020). 10.11586/2021003

13. Folstein, M. F., Folstein, S. E., & McHugh, P. R. (1975). “Mini-mental state”. A practical method for grading the cognitive state of patients for the clinician. Journal of psychiatric research, 12(3), 189–198. 10.1016/0022-3956(75)90026-6

14. García-Casal JA, Franco-Martín M, Perea-Bartolomé MV, Toribio-Guzmán JM, García-Moja C, Goñi-Imiz-coz M, Csipke E. ELECTRONIC DEVICES FOR COGNITIVE IMPAIRMENT SCREENING: A SYSTEMATIC LITERATURE REVIEW. Int J Technol Assess Health Care. 2017 Jan;33(6):654–673. doi: 10.1017/S0266462317000800. Epub 2017 Sep 18. PMID: 28920567.

15. Hemmerich, W. (2018). StatistikGuru: Sensitivität und Spezifität. Retrieved from https://statistikguru.de/lexikon/sensitivitaet-und-spezifitaet.html

16. IBM Corp. Released 2023. IBM SPSS Statistics for Windows, Version 29.0.2.0 Armonk, NY: IBM Corp

17. Isaacs B, Kennie AT. The Set Test as an Aid to the Detection of Dementia in Old People. British Journal of Psychiatry. 1973;123(575):467–470. doi:10.1192/bjp.123.4.467

18. Jack CR Jr, Andrews JS, Beach TG, Buracchio T, Dunn B, Graf A, Hansson O, Ho C, Jagust W, McDade E, Molinuevo JL, Okonkwo OC, Pani L, Rafii MS, Scheltens P, Siemers E, Snyder HM, Sperling R, Teunissen CE, Carrillo MC. Revised criteria for diagnosis and staging of Alzheimer’s disease: Alzheimer’s Association Workgroup. Alzheimers Dement. 2024 Aug;20(8):5143–5169. doi: 10.1002/alz.13859. Epub 2024 Jun 27. PMID: 38934362; PMCID: PMC11350039.

19. Kaplan, E., Goodglass, H., & Weintraub, S. (No year specified.). Boston Naming Test (BNT) [Database record]. APA PsycTests. 10.1037/t27208-000

20. Kuffel, A., Terfehr, K., Uhlmann, C., Schreiner, J., Löwe, B., Spitzer, C., & Wingenfeld, K. (2013). Messwiederholung von Gedächtnistests unter Berücksichtigung der Valenz des Testmaterials: verbales Gedächtnis, Arbeitsgedächtnis und autobiografisches Gedächtnis [Repeated measurement of memory with valenced test items: verbal memory, working memory and autobiographic memory]. Fortschritte der Neurologie-Psychiatrie, 81(7), 390–397. 10.1055/s-0033-1335778

21. Lillig, R., Ophey, A., Schulz, J. B., Reetz, K., Wojtala, J., Storch, A., Liepelt-Scarfone, I., Becker, S., Berg, D., Balzer-Geldsetzer, M., Kassubek, J., Hilker-Roggendorf, R., Witt, K., Mollenhauer, B., Trenkwalder, C., Roeske, S., Wittchen, H. U., Riedel, O., Dodel, R., & Kalbe, E. (2021). A new CERAD total score with equally weighted z-scores and additional executive and non-amnestic “CERAD-Plus” tests enhances cognitive diagnosis in patients with Parkinson’s disease: Evidence from the LANDSCAPE study. Parkinsonism & related disorders, 90, 90–97. 10.1016/j.parkreldis.2021.07.034

22. Mattke S, Tang Y, Hanson M, von Arnim CAF, Frölich L, Grimmer T, Onur OA, Perneczky R, Teipel S, Thyrian JR. Current Capacity for Diagnosing Alzheimer’s Disease in Germany and Implications for Wait Times. J Alzheimers Dis. 2024;101(4):1249–1259. doi: 10.3233/JAD-240728. PMID: 39302379.

23. Mohs RC, Kim Y, Johns CA, Dunn DD, Davis KL. Assessing change in Alzheimer’s disease: Memory and language tests. In Poon LW et al. (Ed.), Handbook for clinical memory assessment of older adults (pp. 149-155). Washington, DC: American Psychological Association, 1986.

24. Morris JC, Mohs RC, Rogers H, Fillenbaum G, Heyman A. Consortium to establish a registry for Alzheimer’s disease (CERAD) clinical and neuropsychological assessment of Alzheimer’s disease. Psychopharmacology Bulletin 1988;24(4):641–52.

25. Müller S, Herde L, Preische O, Zeller A, Heymann P, Robens S, Elbing U, Laske C. Diagnostic value of digital clock drawing test in comparison with CERAD neuropsychological battery total score for discrimination of patients in the early course of Alzheimer’s disease from healthy individuals. Sci Rep. 2019 Mar 5;9(1):3543. doi: 10.1038/s41598-019-40010-0. PMID: 30837580; PMCID: PMC6400894.

26. Randolph, C., Tierney, M. C., Mohr, E., & Chase, T. N. (1998). The Repeatable Battery for the Assessment of Neuropsychological Status (RBANS): preliminary clinical validity. Journal of clinical and experimental neuropsychology, 20(3), 310–319. 10.1076/jcen.20.3.310.823

27. Reissmann, M., Oswald, V., Zank, S., & Tesch-Römer, C. (2022). Digitale Teilhabe in der Hochaltrigkeit. (D80+Kurzberichte, 6). Köln: Bundesministerium für Familie, Senioren, Frauen und Jugend; Universität zu Köln, Cologne Center for Ethics, Rights, Economics, and Social Sciences of Health (ceres); Deutsches Zentrum für Altersfragen.

28. Reitan R. Trail-making test. Arizona: Reitan Neuropsychology Laboratory, 1979.

29. Rosen WG, Mohs RC, Davis KL. A new rating scale for Alzheimer’s disease. Am J Psychiat. 1984;141:1356–1364.

30. Schicktanz, S., Perry, J., Herten, B. et al. Demenzprädiktion als ethische Herausforderung: Stakeholder fordern Beratungsstandards für Deutschland. Nervenarzt 92, 66–68 (2021). 10.1007/s00115-020-00985-y

31. Schmid, N. S., Ehrensperger, M. M., Berres, M., Beck, I. R., & Monsch, A. U. (2014). The Extension of the German CERAD Neuropsychological Assessment Battery with Tests Assessing Subcortical, Executive and Frontal Functions Improves Accuracy in Dementia Diagnosis. Dementia and geriatric cognitive disorders extra, 4(2), 322–334. 10.1159/000357774

32. Seifert, A. Digitale Transformation in den Haushalten älterer Menschen. Z Gerontologie Geriatrie, 55(4), 305–311 (2022). 10.1007/s00391-021-01897-5

33. Seymour, M., Yuan, L. (Ivy), Riemer, K., & Dennis, A. R. (2024). Less Artificial, More Intelligent: Understanding Affinity, Trustworthiness, and Preference for Digital Humans. Information Systems Research. 10.1287/isre.2022.0203

34. Shulman KI, Gold DP, Cohen CA, Zucchero CA. Clock-drawing and dementia in the community a longitudinal study. Int J Geriatr Psychiatry. 1993;8:487–496.

35. Smith, A. (1973). Symbol Digit Modalities Test. Los Angeles, CA: Western Psychological Services.

36. Thurstone, L. L., & Thurstone, T. G. (1962). Primary Mental Abilities. Chicago: University of Chicago Press.

37. Tombaugh T. N. (2004). Trail Making Test A and B: normative data stratified by age and education. Archives of clinical neuropsychology : the official journal of the National Academy of Neuropsychologists, 19(2), 203–214. 10.1016/S0887-6177(03)00039-8

38. Wechsler, D. (1997c). WMS-III Wechsler Memory Scale. San Antonio: Psychological Corporation.

39. Wolfsgruber S, Jessen F, Wiese B, Stein J, Bickel H, Mösch E, Weyerer S, Werle J, Pentzek M, Fuchs A, Köhler M, Bachmann C, Riedel-Heller SG, Scherer M, Maier W, Wagner M; AgeCoDe Study Group. The CERAD neuropsychological assessment battery total score detects and predicts Alzheimer disease dementia with high diagnostic accuracy. Am J Geriatr Psychiatry. 2014 Oct;22(10):1017–28. doi: 10.1016/j.jagp.2012.08.021. Epub 2013 Jun 4. PMID: 23759289.

40. Wurtz, H. M., Manchester, M., Valteau, T., Hanson, H., Scipion, C., Federman, A., & Arias, J. J. (2025). Artificial intelligence as a tool for cognitive impairment screening: Patient perspectives about benefits and limitations. Alzheimer’s & Dementia: Behavior & Socioeconomics of Aging, 1(1), e70005. 10.1002/bsa3.70005

