## Appendix A for "A Self-Administered Digital Test Battery for Early Detection of Cognitive Impairment in Alzheimer’s Disease"

**Table A1**
*Subtest description incorporated in the BraincheX test battery*

| Task Order | Task name | Brief description | Scoring |
|---|---|---|---|
| 1 | Symbol-Digit-Test (SDT) [35] | Assesses processing speed. Participants assign symbols to corresponding numbers. | Total number of correctly assigned symbols in 60 sec. |
| 2 | Visual Memory Test (VMT) | Assesses language-independent visual object recognition. 20 pictures are to be recognized from 35 old and new pictures. | Total number of correctly identified pictures provide a score range of 0-35. |
| 3 | Line-Orientation-Test (LOT) [5] | Assesses visuospatial perception. Participants identify two lines on top page matching two lines on bottom page in 15 sec. per item. | Each correct item scores 1 point (0-10 total). An item scores 0 if one or both lines are incorrect. |
| 4 | Trail-Making-Test (TMT-B) [28] | Assesses visual search speed and executive function (set shifting). Participants connect numbers and letters in alternating order as quickly as possible. Test is terminated after 300 sec. | Total time to complete (max 300 sec.). Errors prevent continuation, impacting performance. Total time is inverted, lower time = better performance, higher time = worse performance. |
| 5 | Clock-Drawing-Test (CDT) [34] | Assesses executive function, visuospatial memory, and motor skills. Participants draw clock numbers with the clock showing the time '10 past 11'. | Scores vary from 1 (perfect clock) to 6 (no reasonable attempt of a clock). A score ≥ 3 is considered impaired. |

**Table A2**
*Subtests of the CERAD-Plus [24]*

| Task Order | Task name | Brief description | Scoring |
|---|---|---|---|
| 1 | Verbal Fluency: Animal Naming [17] | Assesses semantic fluency. Participants name as many animals as possible. | Total number of correctly named animals in 60 sec. |
| 2 | Modified Boston Naming Test [19] | Assesses linguistic ability. Participants are given 10 sec. for each item (15 items) to name them correctly. | Total number of correctly named items provide a score range of 0-15. |
| 3 | Mini-Mental-Status-Examination (MMSE) [13] | Screening tool assesses orientation, concentration, language, and constructive praxis. | Maximum score is 30 points. |
| 4 | Word List Learning [2] | Assesses the ability to remember newly learnt information.10 common nouns (presented for 2 sec.) administered on 3 successive occasions, each time in a different order. The participants read them out loud and then recall them from memory in 90 sec. | Total number of correctly remembered words provide a score range of 0-30. |
| 5 | Figure Copy [29] | Assesses constructional praxis. Participants copy 4 geometric figures increasing in complexity with a pencil (max. 120 sec. per item). Error correction is allowed. | Evaluation of the last attempt or the figure that is indicated by the participants as the best match with the template. Total number of correctly copied figures provide a score range of 0-11. |
| 6 | Word List Recall | Assesses how well the participants remember the 10 words from task 4 in 90 sec. | Total number of correctly remembered words provide a score range of 0-10. |

| | | | |
|---|---|---|---|
| 5 | Word List Recognition [23] | Assesses recognition of the 10 words from task 4, embedded with 10 foils. | Total number of correctly recognized words and correctly rejected new words provide a score range of 0-10. |
| 6 | Figure Recall | Assesses recall of constructional praxis. Participants remember the figures from task 5. | Evaluation of the last attempt or the figure that is indicated by the participants as the best match with the remembered figure. Total number of correctly remembered figures provide a score range of 0-11. |
| 7 | Trail-Making-Test (TMT-A and B) [28] [37\| | Assesses visual search speed (A), and executive function (cognitive flexibility, set shifting, B). Participants connect numbers only (A) or numbers and letters (B) in alternating order as quickly as possible. Test is terminated after 180 (A) and 300 sec. (B). | Total time to complete (max 300 sec.). Errors are assessed indirectly, through additional time spent. Total time is inverted, lower time = better performance, higher time = worse performance. |
| 8 | Phonemic Fluency [36] | Assesses verbal production, semantic memory and speech. Participants name as many words as possible, that begin with the letter 'S'. Not allowed are names of persons, geographical places, numbers or the same word in different forms/ with different endings. | Total number of correctly named words provided in 60 sec. |

**Figure A1**
*Normative Data of BraincheX Subtests*

| Parameters | N | Minimum | Maximum | Mean (M) | Standard Deviation (SD) |
|---|---|---|---|---|---|
| SZT | 56 | 0 | 25 | 13.70 | 7.56 |
| VMT | 54 | 17 | 34 | 28.43 | 4.25 |
| LOT | 56 | 0 | 9 | 4.77 | 2.52 |
| TMT-B | 56 | 53 | 300 | 137.71 | 83.87 |
| Clock Drawing Test | 56 | 1 | 6 | 2.66 | 1.46 |

*Note:* Values calculated based on the healthy control group of the Symbol Digit Test (SZT), Visual Memory Task (VMT), Line Orientation Test (LOT) and Trail Making Test-B (TMT-B).

**Figure A2**
*Coordinates of the ROC-Curve*

| Positive if Greater Than or Equal to | Sensitivity | 1 - Specificity |
|---|---|---|
| -1,97 | ,93 | ,36 |
| -1,77 | ,90 | ,36 |
| -1,51 | ,90 | ,32 |
| -1,26 | ,86 | ,32 |
| -,82 | ,83 | ,32 |
| -,29 | ,79 | ,32 |

Note: Coordinates relevant for specificity and sensitivity were presented in the table. Values are based on the healthy control group and BraincheX total score.
